# A computational approach for measuring sentence information via surprisal: theoretical implications in nonfluent primary progressive aphasia

**DOI:** 10.1101/2022.11.25.22282630

**Authors:** Neguine Rezaii, James Michaelov, Sylvia Josephy-Hernandez, Boyu Ren, Daisy Hochberg, Megan Quimby, Bradford Dickerson

## Abstract

Nonfluent aphasia is a language disorder characterized by simplified sentence structures as well as word-level abnormalities such as a reduced use of verbs and function words. According to the predominant account of the disorder, both structural and word-level features are caused by a core deficit in the processing of syntax. Under this account, however, it remains unclear why nonfluent patients choose semantically richer verbs and may have an intact comprehension of verbs and function words.

Here, we propose and test the hypothesis that the word-level features of nonfluency reflect a process that selects lexically richer words to increase the information content of sentences.

We use a computational linguistic method to measure the information content of sentences in the language of patients with nonfluent primary progressive aphasia (nfvPPA) (n = 36) and healthy controls (n = 133). We measure sentence information using surprisal, a metric calculated by the average probability of occurrence of words in a sentence given their preceding context.

We found that by packaging their structurally simple sentences with lower frequency words, nfvPPA patients produce sentences with similar surprisal as that of healthy speakers. Furthermore, we found that higher sentence surprisal in nfvPPA correlates with a lower function-to-all-word ratio, a lower verb-to-noun ratio, and a higher heavy-to-all-verb ratio.

Surprisal is an effective quantitative index of sentence information. Using surprisal allows for testing an account of nonfluent aphasia that regards word-level features of nonfluency as adaptive rather than defective symptoms, a finding that may entail revisions in therapeutic approaches to nonfluent speech.

## Introduction

Nonfluent aphasia is a language disorder characterized by effortful speech and impaired sentence formation. The sentence impairment can be described at the levels of words as well as structures that determine word relationships.^1,2^ At the structural level, nonfluent patients have difficulty using complex syntactic rules such as embedding one clause into another. At the word level, the impairment is manifested by the use of fewer function words (e.g. pronouns and prepositions), a lower verb to noun ratio, and “heavier” (i.e. semantically richer) verbs than healthy controls. The cause for both levels of impairment has been postulated to be a core deficit in the processing of syntax. That is, deficient syntactic processing would result in the use of simpler structures as well as fewer verbs and function words–the two word types that serve a predominant grammatical role. ^3,4^ This account, however, fails to explain why nonfluent patients tend to use heavy verbs more often, ^5,6^ and may have an intact comprehension of verbs and function words. ^7,8^ While often dismissed by the agrammatic account, these unexplained features might be hinting at a distinct language process that becomes critical for communication in nonfluent aphasia.

Our recent findings based on theories of probabilistic linguistics have raised the possibility of an alternative explanation of the symptoms in nonfluent aphasia. One study showed that when healthy speakers were constrained to produce short sentences of only one to two words, similar word level features of nonfluency emerged in their language.^9^ The extreme production constraint of the experiment helped further accentuate these features. The resemblance between the language of healthy speakers under a production constraint and the typical language of nonfluent patients suggest that this style of word selection might not be the defect, but rather a response to a bottleneck in language production. The work further showed that the words that are commonly dropped by nonfluent patients have a higher frequency of occurrence. According to information theory, outcomes with a high probability of occurrence, such as high frequency words, have a low informational content.^10^ The work thus concluded that nonfluent patients drop the less informative words of a sentence in favor of the more informative ones in response to their deficit in producing complex structures. Further support for this conclusion came from another study that compared the frequency of words and syntactic structures of sentences of patients with nonfluent aphasia.^11^ The work showed that sentences with simpler syntactic structures – as shown by their higher syntax frequency – contain lower frequency words. This tradeoff between the frequencies of words and syntactic rules provides yet another clue to a possible compensatory mechanism in which syntactically impoverished sentences get packaged with more informative words, enabling patients to get their message across. We will refer to the strategy of using more informative words to augment the informational content of a sentence as *lexical condensation*.

The goal of this study is to test such compensatory hypothesis in nonfluent aphasia (Figure 1). If true, this alternative explanation would offer a shift from the prevailing account of agrammatism and might entail a revision in current therapeutic approaches. The compensatory hypothesis can be tested by measuring the informational content of sentences through a metric that factors in both lexical and syntactic information. Through this metric, it becomes possible to explore how these two linguistic elements interact to encompass the information content of a sentence in health or disease. One such metric is *surprisal*, a measure of the likelihood of occurrence of words in a sentence given their preceding context.^12^ Words that have a lower probability of occurrence following a context will be more surprising, and hence have a higher informational content. For example, in the context, “the woman is pouring a ……”, the word *seltzer* has higher surprisal than the word *drink* due to its lower probability of occurrence given the preceding words. Unlike word frequency which reflects the informational content of a word in isolation, surprisal has the critical advantage of being sensitive to its preceding context. Both lexical and syntactic properties of the context will determine the probability of occurrence of the upcoming word,^13^ since not all words fit all structures or strings of words.^14^

**Figure 1.**
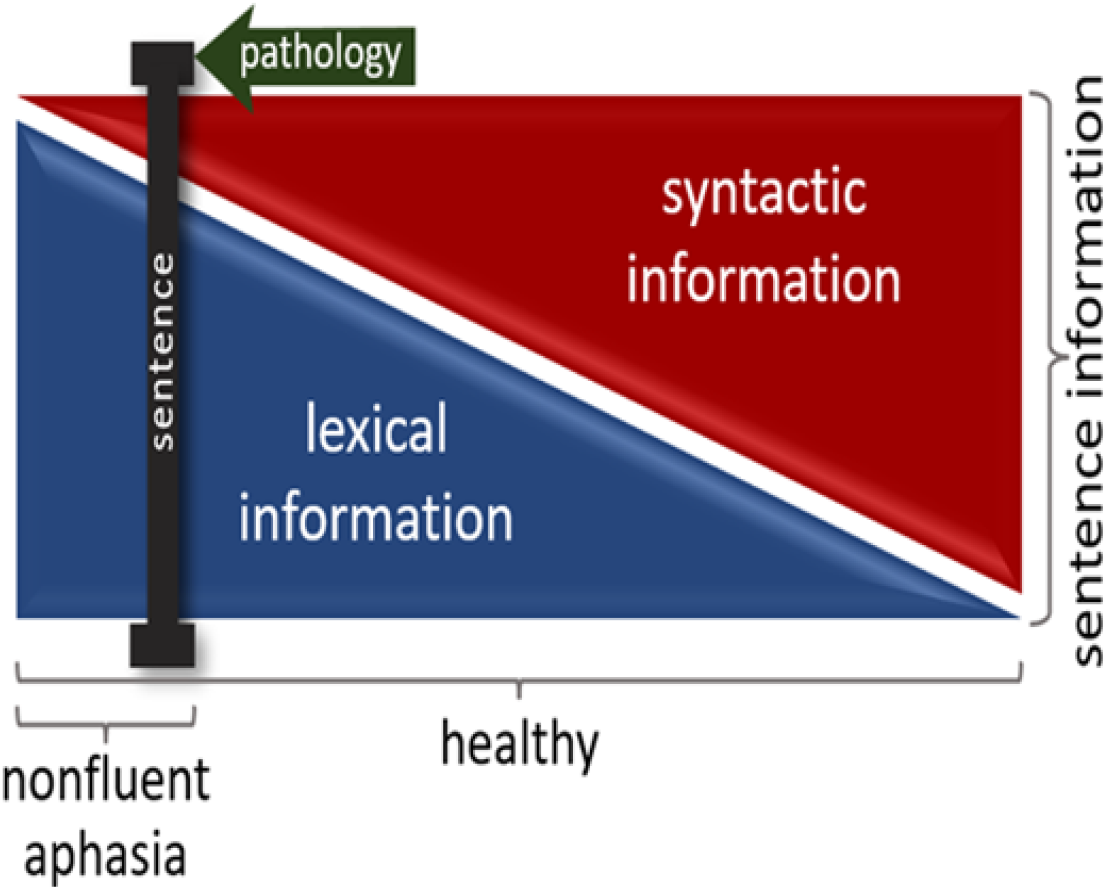
The working hypothesis of the study. Each sentence, represented by the black sliding bar, can be made up of a different share of lexical and syntactic information. In a healthy individual the bar can slide over a wide range of possible combinations of the two sources of information while keeping the sentence information constant. In nfvPPA, the pathological process limits the use of complex syntax, which would push the sliding bar to the left where more informative words need to be selected to convey the intended message. Sentence information is measured by average surprisal, lexical information by average content word frequency, and syntactic information by average syntax frequency.

We calculate surprisal using an automated algorithm, PsychFormers. ^15^ The algorithm is based on Language Models (LMs), computational systems that are designed to assign probabilities to sequences of words. ^16^ The surprisal of a sentence is operationalized as the average surprisal of its words or subwords as recognized by the LM. We will apply this algorithm to language samples collected from patients with the nonfluent variant of primary progressive aphasia (nfvPPA) and healthy controls as they described a picture of a family at a picnic. First, by examining a large sample of language from healthy individuals, we will establish that the surprisal of a sentence is truly a composite metric made up of both lexical and syntactic information. We will measure the information content of words and syntactic rules using their respective frequencies. Next, we will compare the surprisal of spoken sentences from patients with nfvPPA with that of healthy controls. Finding similar surprisal in length-matched sentences produced by the two groups would provide evidence for the proposal that the syntactically simpler sentences of the nonfluent patients become enriched by the use of more informative words. Last, we will repeat the analyses using the written samples of nfvPPA patients and healthy controls describing the same picture. This analysis will help delineate whether the lexical condensation strategy is unique to spoken language or also exists in writing. If observed only in speaking, the compensation strategy might be a response to effortful speech. That is, the cost of articulation is so high that the patients have to resort to short, lexically rich sentences. If the strategy is also present in writing, then the bottleneck is expected to be at an earlier stage in language production that is common to both speaking and writing.

## Methods

### Participants

#### Patients

Thirty-six patients with nfvPPA were recruited from an ongoing longitudinal study being conducted in the Primary Progressive Aphasia Program in the Frontotemporal Disorders Unit of Massachusetts General Hospital (MGH). Of the 36 patients, 34 provided spoken and 30 provided written samples (with 28 patients common to both groups). All patients underwent a standard clinical evaluation comprising a structured history obtained from both patient and informant, comprehensive medical, neurological, and psychiatric history and exams, neuropsychological and speech-language assessments, and a clinical brain MRI scan that was visually inspected for 1) consistency of regional atrophy with a given syndromic diagnosis, and 2) other focal brain lesions or evidence of cerebrovascular disease.^17^ We also include ratings on our scale called the Progressive Aphasia Severity Scale (PASS).^18^ Modeled after the Clinical Dementia Rating Scale (CDR), the PASS uses the clinician’s best judgment, integrating information from the patient’s test performance and a companion’s interview. The PASS includes “boxes” for fluency, syntax, word retrieval and expression, repetition, auditory comprehension, single word comprehension, reading, writing, and functional communication. The PASS Sum-of-Boxes (SoB) is a sum of the box scores. The clinical and demographic information on the patients is shown in Table 1.

**Table 1.** The clinical and demographic information of patients with nfvPPA

| Mean Age<br>(SD) | % Female | % Right-Handed | Mean PASS<br>articulation (SD) | Mean PASS<br>fluency (SD) | Mean PASS<br>SoB (SD) | Mean CDR<br>SoB (SD) |
| --- | --- | --- | --- | --- | --- | --- |
| 67.26<br>(13.58) | 58.3% | 88.9% | 0.97<br>(0.95) | 0.81<br>(0.64) | 6.50<br>(4.47) | 1.60<br>(1.52) |
| PASS = Progressive Aphasia Severity Scale<br>CDR = Clinical Dementia Rating<br>SoB = Sum of Boxes |  |  |  |  |  |  |

#### Healthy controls

A total of 133 native English speakers with no reported history of neurological or acquired/developmental language disorders were recruited to provide spoken (n = 49) and written (n = 84) samples. Thirty-six participants of the spoken cohort were recruited from the Speech and Feeding Disorders Laboratory at the MGH Institute of Health Professions. The rest of healthy participants (13 spoken and 84 written) were recruited from Amazon’s Mechanical Turk (MTurk). Healthy participants had an average age of 50.54 (SD = 16.4) and an average year of education of 15.8 (SD = 1.7). Of all healthy participants, 51.9% were female and 88.9% were right-handed. All statistical analyses that compared two groups were performed on the subsample of healthy controls that matched the age of the participants between the two groups.

### Standard Protocol Approvals, Registrations, and Patient Consents

The recruitment of healthy controls was approved by the Brain Resilience in Aging: Integrated Neuroscience Studies (BRAINS) program at MGH. Obtaining informed consent from all patients as well as the recruitment of all healthy controls were performed in accordance with guidelines established by the Mass General Brigham Healthcare System Institutional Review Boards which govern human subjects research at MGH and specifically approved this study.

#### Language samples

Participants were asked to look at a drawing of a family at a picnic from the Western Aphasia Battery– Revised^19^ and describe it using as many full sentences as they could. The spoken language samples were transcribed into text using Microsoft Dictate application. The transcriptions were then manually checked for accuracy by a research collaborator who was blind to the grouping. Disfluencies of speech such as repetitions and use of fillers, such as “um”, “you know”, etc., were identified per the protocol previously described^11^ and removed from further analyses.

To control for a possible non-linear relationship between sentence length and other language features of a sentence, all comparisons were matched based on sentence length. If there was a significant difference in the sentence length between two groups, we randomly sampled the sentences from each group (without replacement) so that the groups had equal sentence length distribution. In addition, we matched the one-to-two-word sentences that are typical of nfvPPA by including data from a cohort of healthy individuals from MTurk who were asked to describe the same picnic picture using only one-to-two-word sentences. This method allowed us to extend our analyses to the short sentences in nfvPPA.

### Measuring lexical and syntactic features

Word frequency, syntax frequency, noun frequency, verb frequency, function-to-all-word ratio, verb-to-noun ratio, heavy-to-all-verb ratio were automatically extracted using Quantitext, a language toolbox developed in the MGH FTD Unit with the goal of increasing precision while reducing human labor^20^. To determine the part of speech of words, the toolbox uses the automated Stanza Lexicalized Parser.^21^ Nouns, verbs (except *be, have* and *do*), adjectives and adverbs were considered as content words. All other words were classified as function words. We measured function-to-all-word-ratio by dividing the number of function words by the number of all words in a sentence. The verb-to-noun-ratio was measured by dividing the number of verbs by the sum of nouns and verbs in each sentence. The following verbs were classified as light verbs: ‘be’, ‘go’, ‘take’, ‘come’, ‘make’, ‘get’, ‘give’, ‘have’ while excluding auxiliaries from this list.^6^ All other verbs were classified as heavy verbs. Heavy-to-all-verb-ratio was measured by dividing the number of heavy verbs by the total number verbs in a sentence.

To measure word frequency, we used the Switchboard corpus,^22^ which consists of spontaneous telephone conversations averaging 6 minutes in length spoken by over 500 speakers of both sexes from a variety of dialects of American English. We use this corpus to estimate word frequency in spoken English, independently of the patient and control sample. The corpus contains 2,345,269 words. The word frequency of each sentence is calculated by taking the average log frequency of content words within that sentence.

To measure syntax frequency,^11^ we first parsed the sentences in the corpus using Stanza to extract headed syntactic rules. A headed syntactic rule is determined by the head and all its dependents in a dependency parse, whether they occur on the left or right. We applied this method on Switchboard which resulted in 954,616 rules and measured the syntax frequency of each sentence of participants by calculating the average log frequency of the syntactic rules of that sentence based on the Switchboard counts.

### Measuring surprisal

The *surprisal (S)* of a word (w_i_) can be measured by the negative logarithm of its probability given its preceding context w_1_…w_i−1_, as shown in the formula,

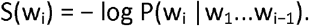

To operationalize the measurement of surprisal, we used PsychFormers,^15^ a tool developed by one of the authors, JM, which allows the surprisal of a word or sequence of words to be calculated using a given transformer language model. Transformer language models have a neural network LM architecture^23^ that has been found to outperform recurrent neural networks,^24^ the previous state-of-the-art architecture, at the standard language modeling task (predicting words from context, see^25^ for review), as well as a range of other tasks.^26,27^ In the present study, we calculate word surprisal using GPT-2^26^ a high-quality transformer language model trained on 40 GB of text data to predict words based on their preceding linguistic context. Specifically, we use the 117 million parameter GPT-2 model made available through the transformers Python library^28^ to calculate the total surprisal of a sentence, which was then divided by the number of tokens in the sentence to get a metric of surprisal normalized by sentence length. A lexical token is a sequence of characters as split up by the language model’s tokenizer, which can be treated as the equivalent of a word in the language model’s vocabulary. In this work, the surprisal of a sentence denotes the normalized sentence surprisal that is the average surprisal of the tokens in a sentence.

### Statistical analyses

For the statistical analyses of this study, we used the R software version 4.1.2. To estimate the smooth but potentially nonlinear relationship between the average surprisal, syntax frequency, and word frequency of a sentence, we used generalized additive models (GAM). GAM is a type of generalized linear model in which the mean of the outcome is a sum of unknown smooth univariate functions of continuous predictors.^29^ Spline functions are popular choices for bases in GAM due to their ability to approximate any smooth function when the number of internal knots is large enough. ^30^ To avoid overfitting and promote generalizability of the fitted model, a smoothness penalty on the spline function is usually used to prevent the model from interpolating. A commonly used class of penalties targets the L2 norm of the derivative of a given order and controls the complexity of the fitted GAM. We use thin plate regression splines^31^ as the basis functions and set the number of internal knots of the spline to be adequately large (e.g. 50). The value of effective degrees of freedom (EDF) formed by the GAM model shows the degree of curvature of the relationship. A value of 1 for EDF is translated as a linear relationship. Values larger than one denote a more complex relationship between the predicting and outcome variables. We used the “gam” function in the “mgcv” package in R to fit the model.^32^ The model parameters were estimated via restricted maximum likelihood (REML) method.^33^ To evaluate the relationship between sentence length, word frequency, and syntax frequency, we combined the language data from spoken and written modalities in healthy controls. Due to sparsity of the distributions of the syntax frequency and word frequency at the two tails which made the gam model prone to noise, we included 99% of the sentences by removing the sparse data at the two tails from both word frequency and syntax frequency distributions. To compare language features at the sentence level across different groups, we used mixed-effects models with subject-specific random intercept via the lme4 package in R. ^34^ To evaluate the relationship between word frequency and other lexical properties of a sentence, we used repeated measures correlation to analyze the common intra-individual association for paired repeated measures. Repeated measures correlation (rmcorr) accounts for intra-participant dependence among repeated observations using analysis of covariance (ANCOVA). By removing measured inter-participant variability, rmcorr provides the best linear fit of the intra-participant relationship between a pair of variables assuming a common slope but varying intercepts across all subjects. We used the rmcorr package from R to perform the analysis.^35^ Bonferroni correction was applied here to adjust for multiple comparisons with the alpha set at 0.01.

### Data availability

The code for measuring surprisal is available at https://github.com/jmichaelov/PsychFormers. Anonymized data not published within this article will be made available by request from any qualified investigator

## Results

### 1. Surprisal is a composite metric made up of lexical and syntactic information

We first evaluated how the surprisal of a sentence is related to the frequency of its content words and syntactic rules in the language samples of healthy individuals, including both written and spoken modalities. We fitted a GAM to the dataset to account for potential non-linearity of the relationship between the variables of interest. We included in the model a subject-specific random intercept, which captures the intra-participant correlation of the repeated measures. We used this model to predict the average surprisal of a sentence from word frequency and syntax frequency.

We found that the surprisal of a sentence could be predicted from both word frequency (*EDF* = 1, *p* < 0.001) and syntax frequency (*EDF* = 2.6, *p* < 0.001) of that sentence. Figure 2 shows the partial effect plots for each smooth term in the GAM that add up to the overall prediction. The value of effective degrees of freedom (EDF) for the word frequency term shows a linear relationship between the surprisal of a sentence and the average frequency of the content words of that sentence. The relationship between the surprisal of a sentence and average syntax frequency is non-linear in a pattern shown in Figure 2.

**Figure 2.**
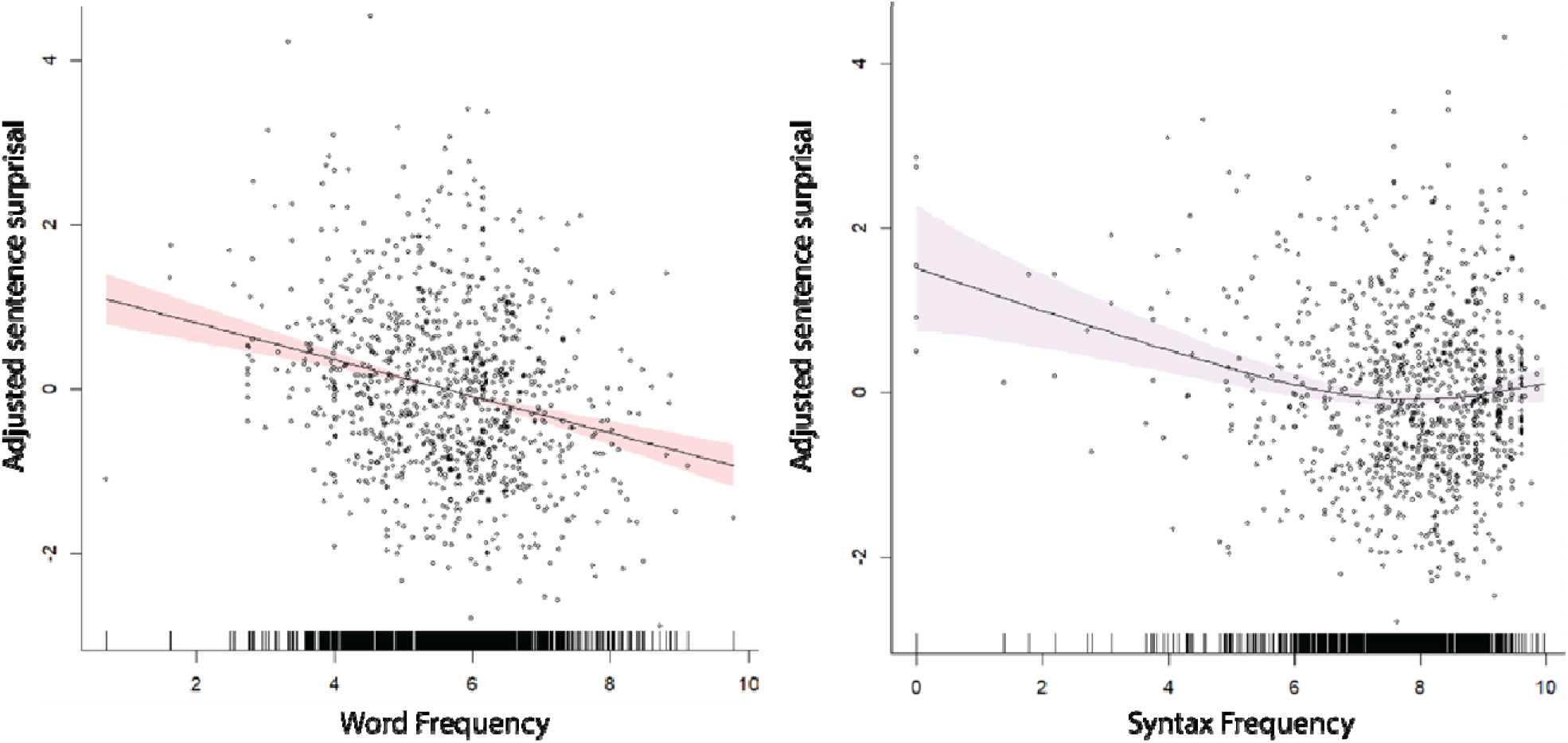
Sentence surprisal is predicted by both word frequency and syntax frequency within each sentence. The graphs show the partial effect plots for each smooth term–word frequency and syntax frequency–in the GAM to illustrate each component of the model predicting sentence surprisal. Shaded zones show 95% confidence intervals around the mean of the effect. In each graph, “adjusted sentence surprisal” represents sentence surprisal adjusted for the other frequency variable (i.e., in the plot showing that word frequency is inversely correlated with adjusted sentence surprisal, sentence surprisal is adjusted for syntax frequency).

### 2. Comparing word frequency, syntax frequency, and surprisal in the spoken and written samples of nfvPPA patients and healthy controls

#### 2.1. By using richer words in their grammatically simpler spoken sentences, nfvPPA patients keep sentence surprisal the same as healthy controls

Fitting a mixed-effects model for sentence length with a fixed effect of subject groups (nfvPPA vs. control) and random intercepts for subjects, we found that patients with nfvPPA produce shorter spoken sentences (mean = 5.57 words, SD = 3.31) than healthy individuals (mean = 8.96 words, 4.49) (β = −3.228, SE = 0.487, t = −6.625, *p* < 0.001). We then examined the difference in other language metrics between nfvPPA patients and healthy control. In light of the statistically significant difference in the distribution of sentence length in the two subject groups as well as the potential nonlinear effects of sentence length on other metrics, we performed the analyses on subsamples of language data from each group with matched distributions of the sentence length. For all the comparisons below, a mixed-effects model was used to predict the variable of interest with group and sentence length as predictors and random intercepts for subjects. The distributions for word frequency, syntax frequency, and surprisal of the two groups are shown in panel A of Figure 3.

**Figure 3.**
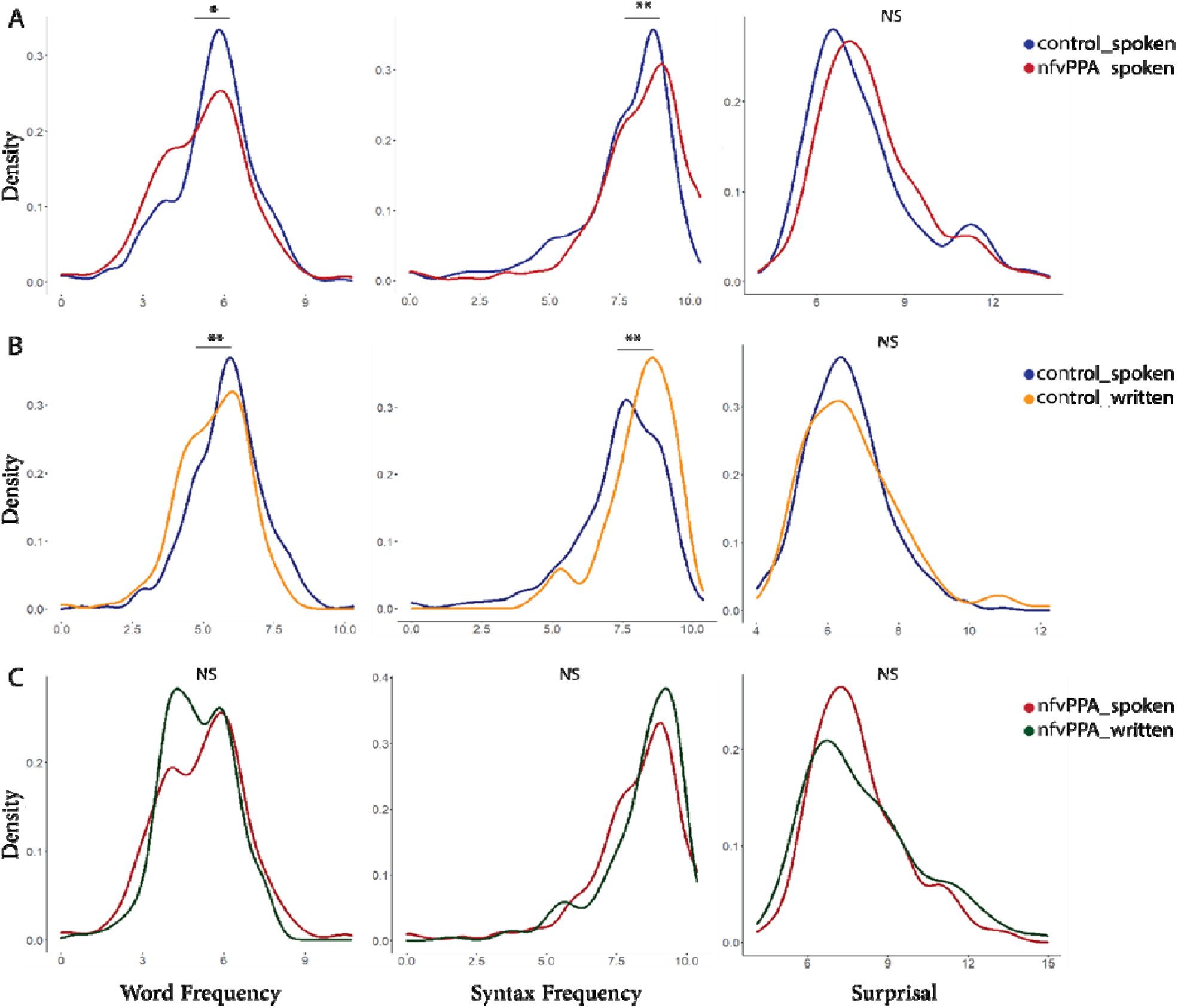
The density graphs of word frequency, syntax frequency, and surprisal at the sentence level across three group pairs. * denotes *p* < 0.05, ** *p* < 0.01, and NS non-significance.

##### Word frequency

Compared to word frequency in the spoken language of healthy individuals (mean = 5.58, SD = 1.54), patients with nfvPPA produced sentences with lower content word frequency (mean = 5.21, SD = 1.70) (β = −0.346, SE = 0.149, t = −2.318, *p* = 0.023).

##### Syntax frequency

Compared to syntax frequency in the spoken language of healthy individuals (mean = 7.63, SD = 1.82), patients with nfvPPA produced syntactic rules of higher frequency (mean = 8.17, SD = 1.74) (β = 0.534, SE = 0.171, t = 3.117, *p* = 0.003).

##### Surprisal

We found no statistical difference between the surprisal of sentences in the spoken samples of healthy controls (mean = 7.59, SD = 1.92) and patients with nfvPPA (mean = 7.81, SD = 1.76) (β = 0.146, SE = 0.209, t = 0.70, *p* = 0.487).

#### 2.2. In healthy individuals, written sentences contain richer words, but simpler syntax, resulting in the same surprisal when compared with spoken language

To compare the sentence length of spoken and written language of healthy individuals describing the same picture, we fit a mixed-effects model to predict sentence length from the modality of language production. We found no statistical difference between the sentence length of spoken (mean = 9.31, SD = 4.74) and written (mean = 8.65, SD = 4.02) language in healthy individuals (β = −0.515, SE = 0.448, t = −1.149, *p* = 0.253). For all the comparisons below, a mixed effects model was used to detect the difference in variables of interest between the two modalities of language production, adjusting for the effect of sentence length. A random intercept is added to capture intra-participant correlations of repeated measures. The distributions for word frequency, syntax frequency, and surprisal of the two groups are shown in panel B of Figure 3.

##### Word frequency

Compared to spoken language (mean = 5.80, SD = 1.27), the written language of healthy individuals contained sentences with lower content word frequency (mean = 5.41, SD = 1.12) (β = −0.458, SE = 0.135, t = −3.389, *p* = 0.001).

##### Syntax frequency

Compared to spoken language (mean = 7.62, SD = 1.55), the written language of healthy individuals contained syntactic structures with a higher frequency of occurrence (mean = 8.12, SD = 1.24) (β = 0.625, SE = 0.190, t = 3.292, *p* = 0.002).

##### Surprisal

We found no statistical difference between the sentence surprisal of spoken (mean = 6.49, SD = 1.18) and written (mean = 6.87, SD = 1.48) modalities in healthy individuals (β = 0.188, SE = 0.151, t = 1.244, *p* = 0.223).

#### 2.3. The spoken and written language samples of patients with nfvPPA show no difference with respect to word frequency, syntax frequency, and surprisal

We repeated the analyses in section 2.2 for patients with nfvPPA. We found no statistical difference between the sentence length of spoken (mean = 5.57, SD = 3.31) and written (mean = 5.54, SD = 2.84) samples in nfvPPA patients (β = 0.232, SE = 0.250, t = 0.926, *p* = 0.355). The distributions for word frequency, syntax frequency, and surprisal of the two groups are shown in panel C of Figure 3.

##### Word frequency

We found no significant difference in content word frequency between spoken (mean = 5.16, SD = 1.63) and written (mean = 5.03, SD = 1.28) language samples of nfvPPA patients (β = −0.145, SE = 0.141, t = −1.025, *p* = 0.306).

##### Syntax frequency

There was no significant difference in syntax frequency between spoken (mean = 8.16, SD = 1.72) and written (mean = 8.40, SD = 1.48) language samples in nfvPPA (β = 0.213, SE = 0.155, t = 1.372, *p* = 0.171). Surprisal. Similarly, we found no significant difference in the sentence surprisal of spoken (mean = 7.90, SD = 1.76) and written (mean = 8.07, SD = 2.15) language in nfvPPA (β = 0.176, SE = 0.141, t = 1.245, *p* = 0.214).

### 3. Word-level features that are correlated with surprisal in the language of nfvPPA patients

Here, we determine which word-level features are correlated with surprisal in the language of patients with nfvPPA at the sentence level after combining spoken and written sentences. We used repeated measures correlation^37^ for this analysis as each participant produced more than one sentence. Figure 4 shows the radar chart of the absolute value of the coefficient of the repeated measures correlations (r_rm_). In the language of patients with nfvPPA, surprisal correlated significantly with function-to-content words (r_rm_ = −0.46, *p* < 0.001), verb-to-noun ratio (r_rm_ = −0.12, *p* = 0.007), heavy-to-all-verb ratio (r_rm_ = 0.15, *p* = 0.006), verb frequency (r_rm_ = 0.24, *p* < 0.001), and noun frequency (r_rm_ = 0.23, *p* < 0.001).

**Figure 4.**
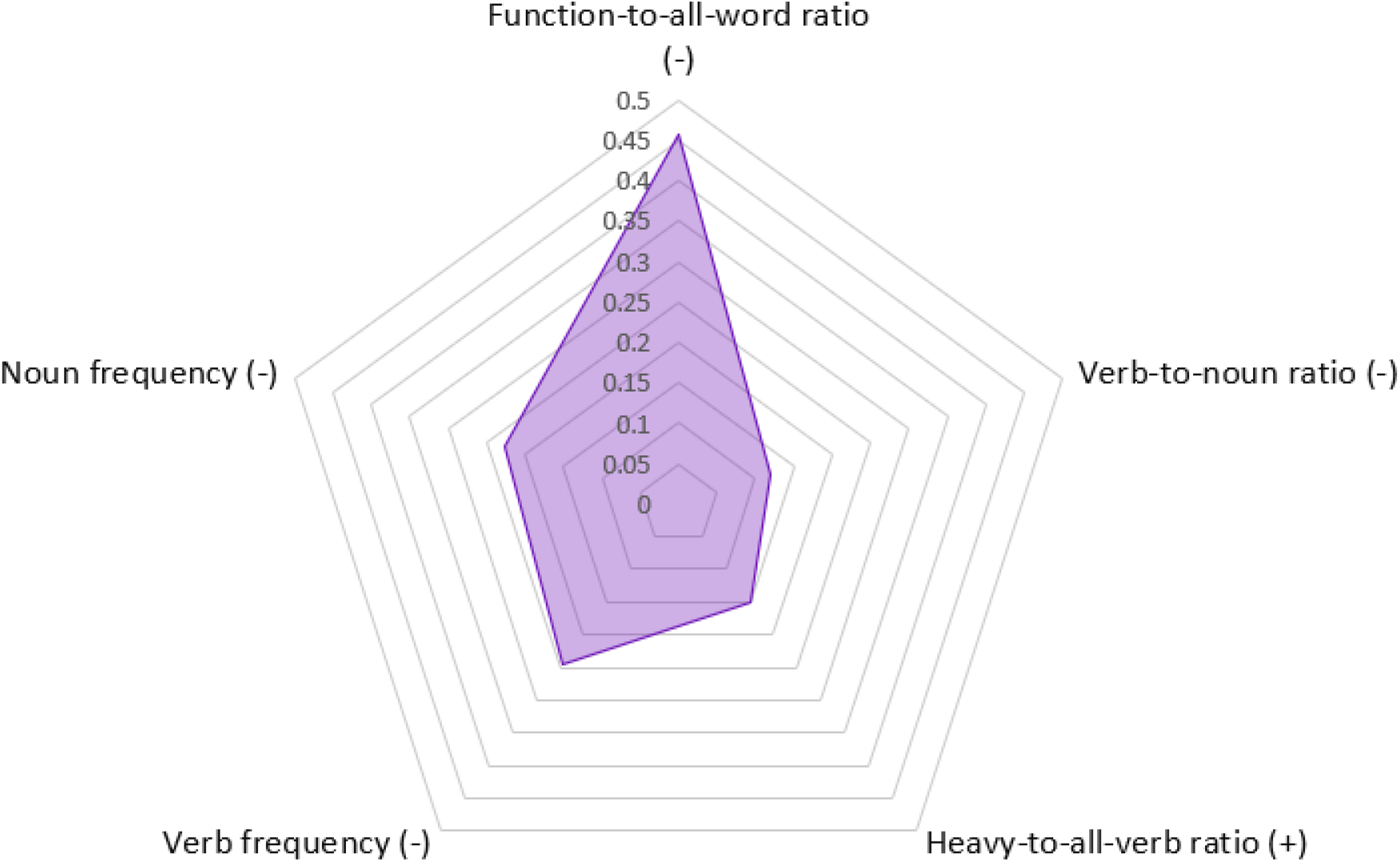
The radar chart shows the absolute value of the coefficient of the repeated measures correlation (r_rm_) between surprisal and various word-level features at the sentence level in the written and spoken samples of nfvPPA patients. The sign in parentheses (+ or -) shows the expected direction of the correlation.

## Discussion

When a brain area is lesioned, it is often a challenge to determine the order of causality among the numerous emerging symptoms. Co-occurrence might lead to the assumption that all symptoms are deficits caused by the lesion. However, given the brain’s tendency to try to restore lost functions, at least partially, it is also possible that some symptoms arise as an adaptive response to the deficit. Although challenging, it is crucial to differentiate adaptive from defective symptoms in order to avoid targeting the wrong symptoms in rehabilitative approaches and to better understand disease mechanism. This challenge exists in the case of the gamut of nonfluent symptoms that arise following a lesion to dominant fronto-insular brain regions. Both word level and structural abnormalities are commonly viewed as defects caused by lesions localized here. In this work, we employed advances in computational linguistics within an information theoretic framework to test the alternative hypothesis that the word level symptoms of nonfluency are in fact an adaptive response to poor sentence structure formation. By measuring sentence information through surprisal, we showed that patients with nfvPPA can produce sentences with the same amount of information per word as healthy controls. To achieve the same level of surprisal, the syntactically impoverished sentences of nonfluent patients get packaged with more informative words, as measured by their lower frequency, through lexical condensation. Based on previous studies, surprisal as calculated by LMs correlates well with behavioral measures of processing difficulty such as reading time ^36,37^ and neural measures such as the N400. ^38,39^ For the purposes of this study, surprisal was particularly suitable as it factors in both lexical and syntactic information allowing us to probe variations in surprisal as a collective representation of these two sources of information in sentences. We further showed that word-level features of nonfluency such as low function-to-all-word ratio, low verb-to-noun ratio, and more heavy-to-all-verb ratio are correlated with higher surprisal.

This evidence for lexical condensation in nonfluent aphasia revives a series of accounts based on compensation from the past century. According to ideas regarding “economy of effort”,^40,41^ the intensive effort required to articulate speech caused by the lesion force nonfluent patients to plan short strings of only essential words. Later, adaptation theory considered sentence processing in nonfluent aphasia to be so slow that the sentence elements would disappear from memory before the syntactic operation is completed. ^42,43^ As a result of this slowed process, nonfluent patients limit the number of sentence elements that need to be retained in memory by using the most essential words. Although theoretically plausible, in the pre-computational linguistics era, these proposals lacked rigorous methods to test their claims.

In healthy human language production, lexical and syntactic processing are intricately intertwined. ^15,49^ Lexical identification and structural scaffolding are two processes that work in close coordination to ensure that the intended message gets across. A bottleneck in the flow of information in one domain, be it lexical or syntactic, would instigate efforts to recast the message through adjustments in the other domain. We posit that the interactivity between syntax and the lexicon enables nfvPPA patients to readily adapt to their difficulty making complex structures. Over time, however, the adjustment to the use of richer words may gradually influence the ways in which these patients formulate thought. ^44^

The dynamic balance of lexical and syntactic information in healthy individuals is also evident in the comparison of speaking and writing. Even though the message across both modalities was kept constant through the same picture description task, we found that written samples contained lower frequency words compared to speaking. This finding is consistent with the established literature that lexical information is richer in writing as measured through various metrics such as higher lexical diversity, ^45,46^ more attributive adjectives, ^46^ and higher lexical density (as measured by the number of content words over either total number of words or clauses). ^47,48^ We further showed that the content words of lower frequency in writing were embedded in syntactic structures of higher frequency. The finding suggests that the writing mode, which is often under less processing demands than speaking, allows for accessing lower frequency words, but at the cost of using simpler structures. ^49^ The net effect of this balance is the same amount of surprisal between written and spoken sentences.

Unlike healthy individuals, nfvPPA patients did not show the flexibility of changing the share of lexical and syntactic information across speaking and writing. We found no difference in word frequency, syntax frequency, and surprisal between the two modalities in nfvPPA patients. This rigidity is likely due to the fixed deficit in using syntactically complex sentences that limits the choice of syntactic rules to only simple structures. As a result, language production in nfvPPA patients might already be at the maximum capacity for using low frequency words.^50^ Lastly, the comparison between speaking and writing in nfvPPA makes it possible to further probe the functional locus of the bottleneck in language production. Our findings rule out the possibility for the articulatory stage to be the limiting factor as the same lexical and syntactic properties were found in the written samples. The functional locus of the bottleneck should thus be explored at an earlier stage of production that is common to both modalities. Future work is needed to explore the stages of high-level motor planning, retrieving and assembling sentence elements, or message conceptualization as potential candidates.

## Data Availability

All codes are available at https://github.com/jmichaelov/PsychFormers. Access to patients data will be granted through a request to Dr. Bradford Dickerson

## Acknowledgements

This research was supported by NIH grants R01 DC014296, R21 DC019567, R21 AG073744, and by the Tommy Rickles Chair in Primary Progressive Aphasia Research. We thank Jordan Green and Claire Cordella for the provision of healthy control data from the Speech and Feeding Disorders Laboratory at the MGH Institute of Health Professions. We thank Arash Afraz for comments on this work. The authors would like to express particular appreciation to the participants in this study and their family members, without whom this research would not have been possible.

